# The associations of child screen time with psychiatric problems: the role of genetic confounding

**DOI:** 10.1101/2023.03.21.23286931

**Authors:** Yingzhe Zhang, Karmel W Choi, Scott W. Delaney, Tian Ge, Jean-Baptiste Pingault, Henning Tiemeier

**Affiliations:** Department of Epidemiology, Harvard T.H. Chan School of Public Health, Boston, MA, USA; Center for Precision Psychiatry, Department of Psychiatry, Massachusetts General Hospital, Boston, MA, USA; Psychiatric & Neurodevelopmental Genetics Unit, Center for Genomic Medicine, Massachusetts General Hospital, Boston, MA, USA; Department of Environmental Health, Harvard T.H. Chan School of Public Health, Boston, MA, USA; Department of Clinical, Educational and Health Psychology, University College London, London, United Kingdom; Social, Genetic, and Developmental Psychiatry Centre, King’s College London, London, United Kingdom; Department of Social and Behavioral Sciences, Harvard T.H. Chan School of Public Health, Boston, MA, USA

## Abstract

**Importance:** Children’s exposure to screen time has been associated with poor mental health outcomes, yet the role of genetic factors in this association remains largely unknown.

**Objective:** We examined (1) the longitudinal phenotypic association between child screen time and mental health outcomes and (2) the potential genetic confounding of this association. We hypothesized that genetics partially account for observed phenotypic associations.

**Design:** Longitudinal (baseline and one-year follow-up) population-based cohort.

**Setting:** Adolescent Brain Cognitive Development, 21 sites in the United States.

**Participants:** This study included 4,262 children of genetically assigned European ancestry with mean age 9.9 years [SD = 0.6 years], 46.8% female.

**Exposure:** Children’s daily screen time (in hours) was assessed both by child-report and parent-report questionnaires at baseline.

**Main Outcomes and Measures:** Child psychiatric problems, specifically attention and internalizing problems, were measured with the parent-rated Child Behavior Checklist at the one-year follow-up. We used Genetic sensitivity analyses (Gsens), based on structural equation models using polygenic risk scores (PRS) of both exposure and outcomes, and either single nucleotide polymorphism (SNP)-based heritability or twin-based heritability to estimate genetic confounding of associations between child screen time and attention or internalizing problems, separately.

**Results:** We found that child screen time was positively associated with the different psychiatric problems. Further, the television time PRS was associated with child screen time (β=0.18 SD, 95% CI: 0.14, 0.23); the ADHD PRS was associated with attention problems (β=0.13 SD, 95% CI: 0.10, 0.16); and the depression PRS was associated with internalizing problems (β=0.10 SD, 95% CI: 0.07, 0.13). These PRSs were associated with cross-traits, suggesting genetic confounding. Using PRSs and SNP-based heritability, we estimated that genetic confounding entirely accounts for the association between child screen time and attention problems, and moderately (42.7%) accounts for the association between child screen time and internalizing problems. When PRSs and twin-based heritability estimates were used, genetic confounding fully explained both associations.

**Conclusions and Relevance:** Genetic confounding may explain a substantial part of the associations between child screen time and psychiatric problems. Potential interventions to reduce screen time could be less effective in reducing psychiatric problems than previously hypothesized.

## Introduction

The potential negative effects of screen time on child mental health are widely discussed. In a recent meta-analysis of 87 studies, greater duration of screen time was associated with small increases in child internalizing problems (r=0.07; 95% CI, 0.05-0.08).^1^ Studies also suggested screen time is associated with more childhood attention problems,^2^ while more social media and television use is related to adolescent depressive symptoms.^3-6^ Similarly, a large-scale population-based study in United States reported that more than 4 hours per day of screen time use among children and adolescents was associated with increased mood and attention problems.^7^ Although some inconsistent results have emerged,^8,9^ excessive screen time is widely recognized to be associated with child psychiatric problems.

Child psychiatric problems, including attention and internalizing problems, are influenced by genetics.^10,11^ Scientists have also begun to investigate how genes influence behavioral traits, including time spent watching television,^12,13^ suggesting that genetics may affect screen time use. Although phenotypic associations between screen time and psychiatric problems have been widely studied, the potential role of genetics in these associations remains largely unknown. As both screen time and psychiatric problems may be influenced by genes, genetic confounding may generate spurious or non-causal associations between child screen time and psychiatric problems.^14,15^ Given interest in interventions to reduce screen time and increase child wellbeing,^16^ evaluating potentially confounding genetic effects in these relationships has important public health implications.

Prior studies considering genetic confounding in other contexts have often adjusted their analyses for polygenic risk scores (PRS).^17,18^ Because PRS is based on imperfectly measured additive effects of common variants, it can be construed as a noisy measure of heritability that generally explains little variance of any given psychiatric trait.^19,20^ Thus, adjusting models for PRSs likely underestimates the confounding arising from genetic factors. To correct the measurement error of using PRS to represent genetic factor, Pingault et al, 2021, proposed a new genetic sensitivity analysis method called “Gsens” to use information from both PRSs and more comprehensive heritability estimates.^21^ In this framework, the overall phenotypic associations are divided into (1) genetic confounding effects and (2) the residual association that excludes genetic confounding. Specifically, in the Gsens model, either single nucleotide polymorphism (SNP)-based heritability (hSNP) or twin-based heritability (htwin) estimates can be used in structural equation models (Figure S1) to account for possible PRS measurement error.

In the model (Figure S1), the polygenic score G measures the underlying genetic factor, which includes measurement error. The latent variable G* captures the heritability of the corresponding trait under the hSNP or htwin scenario. The hSNP captures common genetic variations, but not rare variants,^22^ insertions, or deletions that may partially account for the heritability of Attention Deficit Hyperactivity Disorder (ADHD) and depression.^23,24^ Thus, analyses using hSNP provide a lower (i.e., possibly underestimated) bound of genetic confounding. In contrast, htwin may overestimate heritability due to genetic interactions,^25^ gene-environment interactions,^26^ or by violating the equal environment assumption in twin studies.^27^ Thus, analyses using htwin provide an upper-bound estimate of genetic confounding. Hence, Gsens provides a range for potential genetic confounding effects under either hSNP or htwin scenarios.

Using Gsens, we aimed to assess the extent of genetic confounding in associations between child screen time and attention or internalizing problems in a large population-based cohort. We hypothesized that genetic factors combining both polygenic and different heritability estimates would account for a substantial part of these associations.

## Methods Participants

We used data from the Adolescent Brain Cognitive Development (ABCD Release 4.0) Study of children aged 9-11 years across the United States.^28,29^ Because PRSs were derived from genome-wide association studies (GWAS) in European ancestry populations, we included only genetically unrelated children with genetically identified European ancestry. We compared descriptive statistics between included samples versus self-identified European participants without genetic data for a non-response analysis.

### Genotyping and PRSs

Uban et al, 2018, provided detailed information about genotyping in ABCD.^30^ We used release ABCD 2.0.1 genotype data. After quality control and imputation, we extracted 6,833,710 genetic variants with minor allele frequency >0.01, imputation info score >0.8, SNP-level call rate >0.98, and Hardy–Weinberg equilibrium p >1e-10 (see Supplemental Methods).

To calculate PRSs, we chose large-sample GWASs of phenotypes corresponding to the ABCD child measures (see below). Although these GWASs were conducted in adult samples, previous research suggested a robust overlap between salient genetic factors in children and adults.^31^ We computed genome-wide PRSs using statistics from the following GWAS data: (i) leisure television watching time (n=365,236 individuals);^12^ (ii) ADHD (n=55,374 individuals);^32^ and (iii) major depression (n=500,199 individuals).^33^ PRSs for all traits were computed using PRS-CS software, a Bayesian scoring method which places a continuous shrinkage (CS) prior on SNP effect sizes.^34^

### Child screen time

Participating children completed the 14-question Screen Time Questionnaire (STQ) at baseline, providing self-report measures of screen time use (see Supplemental Methods). The children’s caregiver also completed a shorter version of the STQ about their child’s total screen use that did not measure specific screen time subtypes. Because we used parent-reported child psychiatric outcome data (described below), child-reported screen time was used as the primary exposure to avoid shared-reporter bias while the parent-reported measure was used in sensitivity analyses.

### Child psychiatric problems

Parents completed the Achenbach Child Behavior Checklist (CBCL) 6/18 at one-year follow-up.^35^ We assessed attention problems using the attention problem subscale (10 items) and internalizing problems using the combined anxious/depressed, somatic complaints, and withdrawn/depressed subscales (32 items).

### Covariates

Age, sex, and study site were included as confounders between child screen time and psychiatric problems. Family income, highest parental education, and maternal psychopathology were considered as additional potential confounders (see Supplemental Methods). We also adjusted our models for the top 10 principal components (PCs) to account for residual confounding by genetic ancestry.

### Heritability

We assessed hSNP of screen time, attention problems, and internalizing problems using GCTA-GREML software,^36^ adjusting for age, sex, site, and top 10 PCs. We used rank-based, normality-transformed screen time and log-transformed psychiatric problem scores because of the normality distribution assumption when estimating the heritability. We calculated htwin using 216 pairs of European monozygotic twins and 333 pairs of dizygotic twins from the ABCD study using identity by descent segments^37^ (see Supplemental Methods).

### Statistical Analysis

We calculated the descriptive statistics for included participants. In primary analyses, we first examined associations between child-reported screen time and parent-reported child attention problems or internalizing problems using linear regressions. Second, using linear regressions again, we quantified associations of the PRSs (television time, ADHD, depression) with screen time and psychiatric problems.

Third, we used the Gsens framework to quantify genetic confounding for the associations between screen time and attention and internalizing problems. We fit three sets of structural equation models. The first set used PRSs for the exposure and outcomes to adjust for genetic confounding in a simplistic way. In the second set, we modeled both hSNP and the PRSs to produce a lower-bound estimate of genetic confounding. In the third set, we modeled both htwin and PRSs to delineate the upper bound of genetic confounding. These models used standardized correlations adjusting for sex, age, study site, and PCs. Because the observed PRS for screen time was quite predictive, our models used genetic information of both the exposure and the outcomes. For comparison, we also fit the models using only PRSs for the outcomes.

In sensitivity analyses, we additionally adjusted our models for family income and highest parental education. Separately, we compared associations between screen time and psychiatric problems using screen time data from children versus parents. We also explored how the magnitude of genetic confounding may vary given different estimates of heritability. For this, we conducted Gsens analyses across an hSNP range between 0.01 to 0.3 for psychiatric problems and 0.077 (from our sample) to 0.16 (from prior research) for screen time.^12^

In our analyses, all PRSs, child-reported and parent-reported screen time, and child psychiatric problems were standardized to mean 0 and standard deviation (SD) 1 to facilitate interpretation and comparisons. Under all heritability scenarios, the direction of the effect of screen time on child psychiatric problems was constrained to be positive based on the main phenotypic associations (i.e., the minimum association was null rather than an unlikely protective effect of screen time). We used R v4.0.3 for all analyses.

## Results

### Sample characteristics

Our sample included 4,262 children (47% female) with mean baseline age 9.9 (SD = 0.6) years (Table 1). On average, children reported 3.2 (SD = 2.6) hours of daily screen time. Using a shorter questionnaire capturing only rough screen use hours, parents reported 1.2 (SD = 0.6) hours per day of screen time for their children. Mean attention and internalizing problem scores at one-year follow-up were 2.9 (SD=3.4) and 5.4 (SD=5.6) points, respectively. See Table S1 for the non-response analysis.

**Table 1:** Demographic distribution of European unrelated individuals.

| | Mean ± SD / N (%) |
| --- | --- |
| N | 4262 |
| Age, year | 9.93 ± 0.62 |
| Sex |  |
| Female, % | 1993 (46.8) |
| Male, % | 2269 (53.2) |
| Family annual income |  |
| Less than \$50,000, % | 500 (11.7) |
| \$50,000 - \$100,000, % | 1337 (31.4) |
| More than \$100,000, % | 2425 (56.9) |
| Highest parent education level (degree) |  |
| High School graduation, % | 939 (22.0) |
| Bachelor's Degree, % | 1323 (31.0) |
| Graduate Degree or above, % | 2000 (46.9) |
| Maternal psychopathology, score | 20.81 ± 16.54 |
| Screen time (child-reported) at baseline, hour | 3.18 ± 2.55 |
| Screen time (parent-reported) at baseline*, hour | 1.16 ± 0.60 |
| Attention problems at year 1 follow-up, score | 2.88 ± 3.42 |
| Internalizing problems at year 1 follow-up, score | 5.40 ± 5.59 |

\*Parent-reported screen time questionnaire was less detailed than the child reported one, only asking for overall screen time in average.

### Heritability estimates of screen time and child psychiatric problems

Using GCTA-GREML, the covariate-adjusted hSNP estimates for child-reported screen time were 0.08 (SE=0.08) and 0.06 (SE=0.08) for parent-report measures (Table 2). The covariate-adjusted hSNP estimates were 0.18 (SE=0.08) for attention problem scores and 0.07 (SE=0.08) for internalizing problem scores. Next, based on 216 monozygotic and 333 dizygotic twin pairs, the htwin estimates were 0.57 for child-reported screen time, 0.88 for attention problems, and 0.48 for internalizing problems. We did not use the htwin estimate for parent-reported screen time because many parents reported the same amount of time for each twin, leading to an underestimated htwin.

**Table 2:** Heritability estimates and corresponding sample sizes.

|  | SNP-based N (ABCD) | h<sub>SNP</sub> estimated (ABCD) | SNP-based N (literature) | h<sub>SNP</sub> estimated (literature) | Twin-based N in pairs, (ABCD) | h<sub>twin</sub> estimated (ABCD) | Twin-based N in pairs, (literature) | h<sub>twin</sub> estimated (literature) |
| --- | --- | --- | --- | --- | --- | --- | --- | --- |
| Attention problems | 4,314 | 0.182 (SE=0.081) | 55,374 | 0.216<sup>32,a</sup> | 214 MZ, 333 DZ | 0.88 | 17,026 MZ, 42,488 DZ | 0.88<sup>47,a</sup> |
| Internalizing problems |  | 0.070 (SE=0.077) | 64,641 | 0.017<sup>48,b</sup> / 0.089<sup>33,c</sup> |  | 0.48 | 4,367<sup>*</sup> | 0.42<sup>49,d</sup> |
| Child-reported screen time | 4,303 | 0.077 (SE=0.079) | 422,218 | 0.161<sup>12,e</sup> | 216 MZ, 328 DZ | 0.58 | 1,961 MZ, 3,220 DZ | 0.37<sup>50,f</sup> |
| Parent-reported screen time | 4,273 | 0.059 (SE=0.080) |  |  | NA | NA | NA | NA |

Note: The parent-reported screen time values in twins were highly correlated that twin-based heritability cannot be reliable based on the given information. Screen time was transformed by rank-based normalization and attention problems and internalizing problems phenotypes used the raw scores from the Child Behavioral Checklist. MZ: monozygotic twins. DZ: dizygotic twins.
<sup>a</sup> Clinically diagnosed ADHD cases and controls were used in the analysis.
<sup>b</sup> Self-reported internalizing problems of children aged 3 to 18 were used as the phenotype.
<sup>c</sup> Adult depression measured by broad self-reported questions was used as phenotype.
<sup>d</sup> The Moods and Feelings Questionnaire (MFQ) reported by parents at 12-year-old was used as the phenotype.
<sup>e</sup> Television time in adults was used as the phenotype.
<sup>f</sup> Screen time of entertainment among 16-year-old adolescents was used as the phenotype.
<sup>\*</sup> The original paper did not provide specific numbers in DZ and MZ.

### Screen time and child psychiatric problems

In fully adjusted models, each additional standard deviation of child-reported screen time was associated with a 0.10-SD (95% CI = 0.07, 0.13) increase in attention problem score and a 0.03-SD (95% CI = 0.003, 0.06) increase in internalizing problem score. See Figure 1. Similarly, each additional standard deviation of parent-reported screen time was associated with a 0.10-SD (95% CI = 0.07, 0.13) increase in child attention problem score in a model adjusting for family income and highest parental education, but this estimate was attenuated and non-significant when additionally adjusting for maternal psychopathology (β = 0.02, 95% CI = -0.01, 0.05).

**Figure 1:**
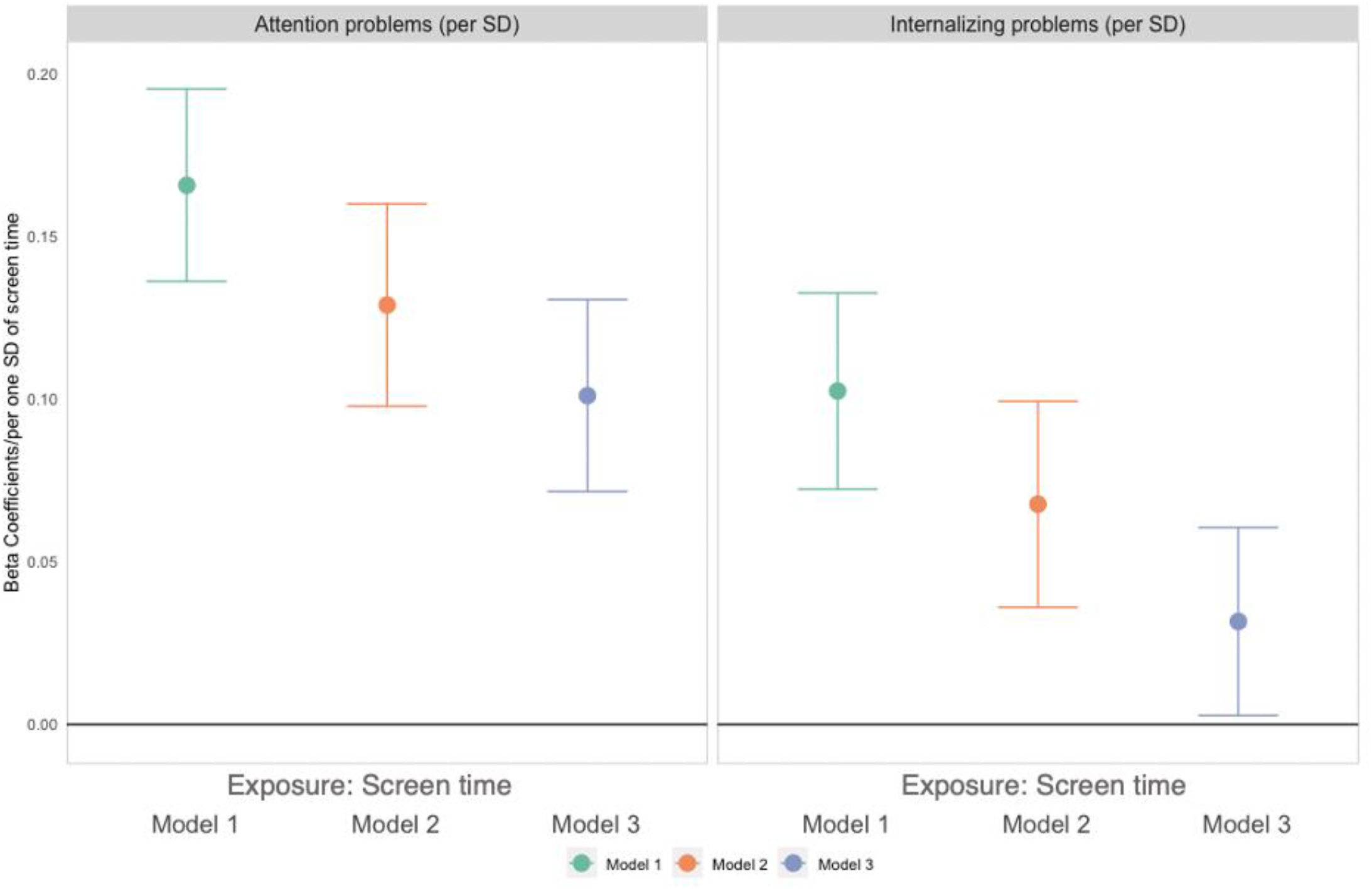
Associations of child reported screen time with parent reported child attention and internalizing problems (N=4262). Model 1 adjusted for sex and age. Model 2 adjusted for sex, age, family income and highest parental education. Model 3 additionally adjusted for maternal psychopathology. Note: Screen time, attention and internalizing problems scores were standardized to mean 0 and standard deviation 1.

Meanwhile, each additional standard deviation of parent-reported screen time was associated with a 0.05-SD (95% CI = 0.02, 0.08) increase in child internalizing problem score in a fully adjusted model (Figure S3).

### Genetic risk scores and screen time

A 1-SD higher television time PRS was associated with a 0.18-SD longer child-report screen time (95% CI = 0.14, 0.23). The PRSs of ADHD and depression were, to a lesser extent, positively associated with screen time: β = 0.14-SD (95% CI = 0.11, 0.17) and β = 0.07-SD (95% CI = 0.04, 0.10), respectively (Figure 2, left panel). For comparison, parent-reported child screen time was also associated with all three PRSs but with smaller magnitudes (Figure S4).

**Figure 2:**
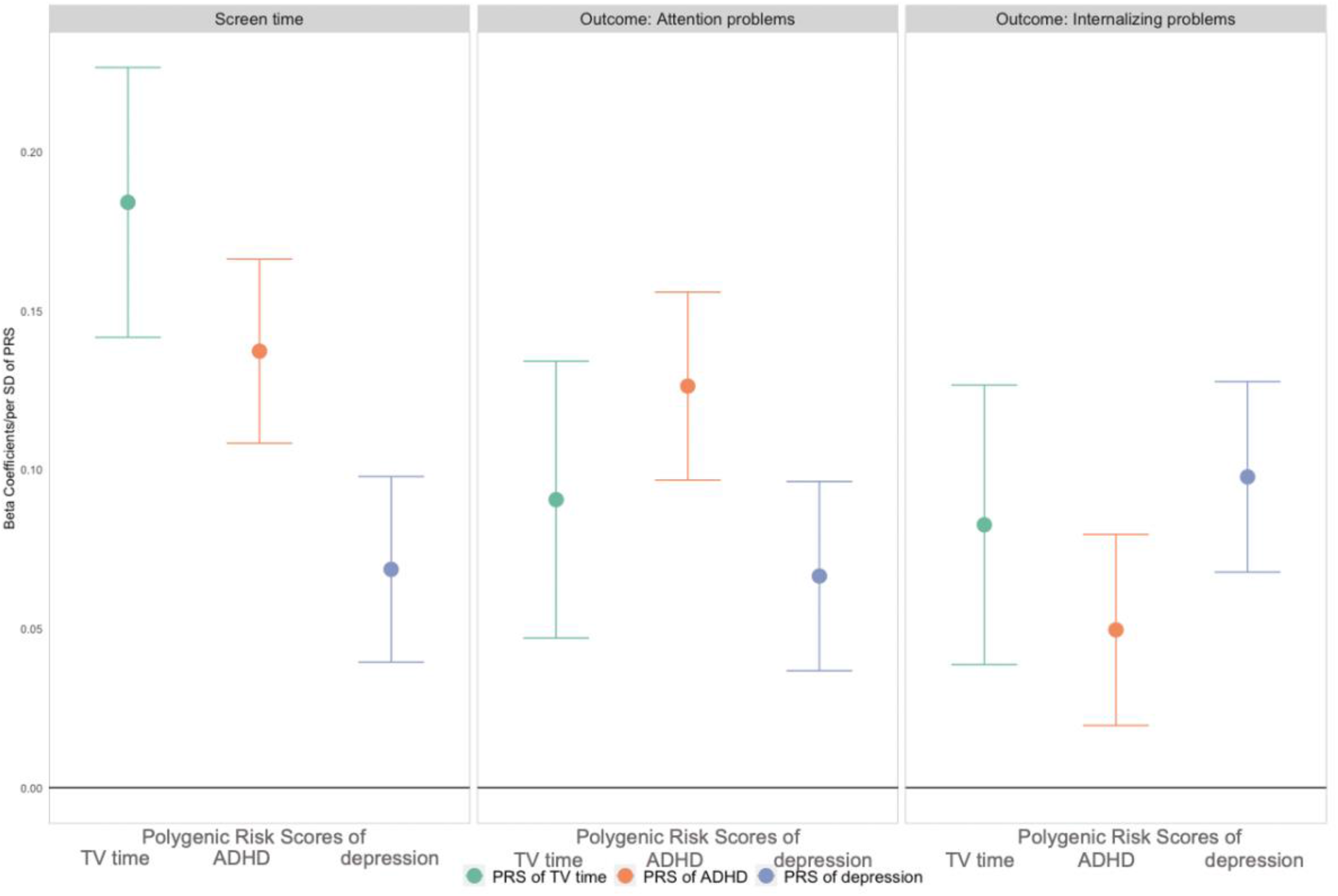
Associations of polygenic scores with screen time, attention and internalizing problems (N=4262). Associations adjusted for age, sex, site, and top 10 PCs. Noted: Polygenic risk scores, screen time, attention problems and internalizing problems were standardized to mean 0 and standard deviation 1.

### Genetic risk scores and psychiatric problems

A 1-SD higher ADHD PRS was related to a 0.13-SD (95% CI = 0.10, 0.16) higher attention problem score. The PRS of television and depression were, to a lesser extent, positively associated with attention problem: β = 0.09-SD (95% CI = 0.05, 0.13) and β = 0.07-SD (95% CI = 0.04, 0.10) (Figure 2, middle panel). Likewise, all PRSs were positively related to internalizing problems (Figure 2, right panel; Table S3).

### Genetic confounding effect

In a model adjusted for both screen time and attention problem PRSs (not using hSNP or htwin), we found a small but significant genetic confounding effect (10.4%), indicating that roughly 10% of the phenotypic association between screen time and attention problems was explained by the genetic confounding (Figure 3A, left panel). However, when using the hSNP estimates for screen time and attention problems, the magnitude of genetic confounding was much larger, such that association between screen time and attention problems was no longer statistically significant (Figure 3A, middle panel). Finally, when using htwin instead of hSNP, our results suggested that genetic confounding may entirely account for the association between child screen time and attention problems (Figure 3A, right panel).

**Figure 3:**
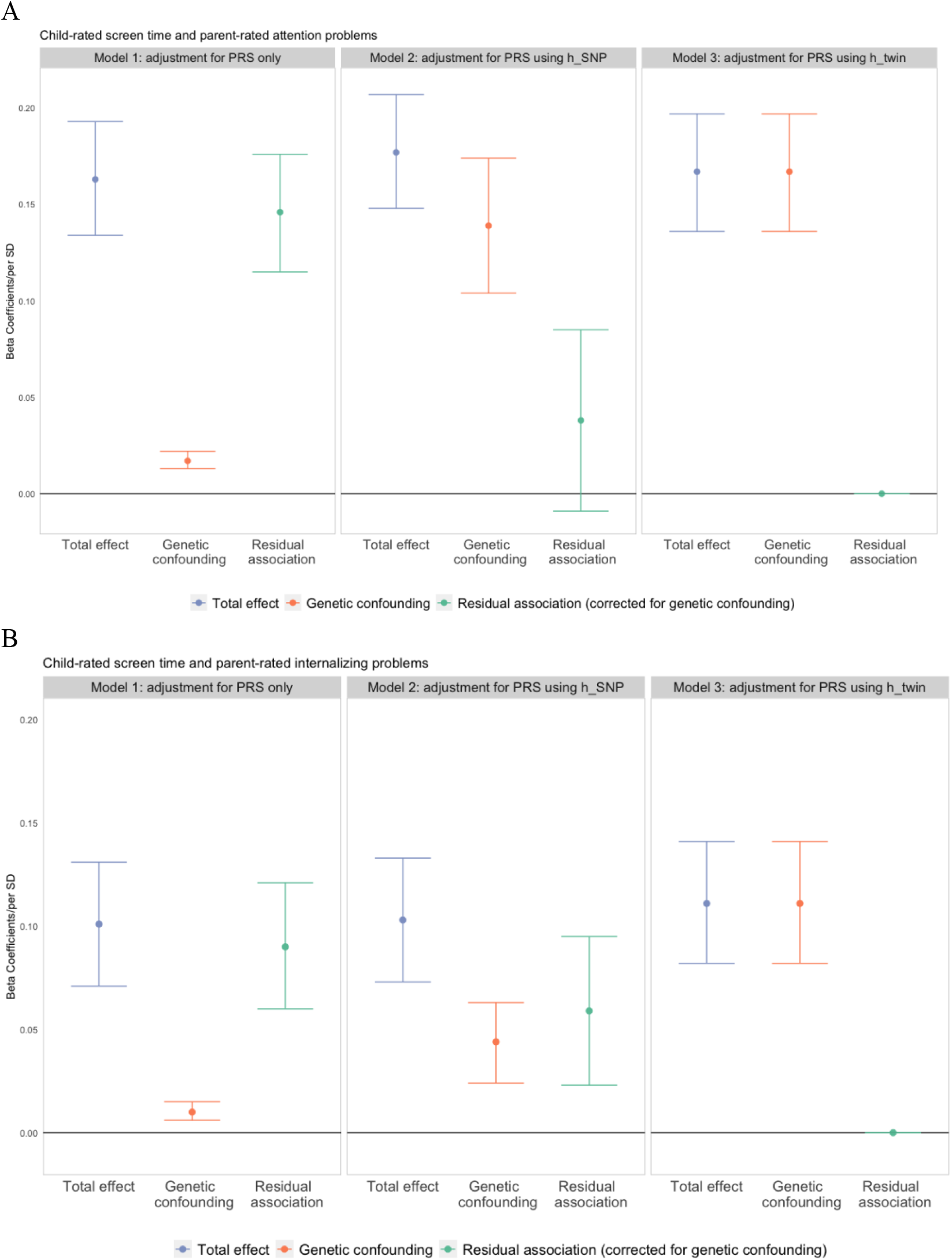
Association of screen time with attention problems and internalizing problems: with different adjustments for genetic confounding (N=4262). Model 1: adjusted for polygenic risk scores for both exposure and outcomes only. Model 2: adjusted for polygenic risk scores using SNP-based heritability (hSNP) for both exposure and outcomes. Model 3: adjusted for polygenic risk scores using twin-based heritability (htwin) for both exposure and outcomes.

Models of genetic confounding of the screen time-internalizing problem relationship revealed similar patterns. Adjusting for PRSs only, we found a significant 10.0% genetic confounding effect for the association between screen time and internalizing problems (Figure 3B, left panel). Using the hSNP estimates, we observed a larger genetic confounding effect of 42.7% (Figure 3B, middle panel). Lastly, using the htwin estimates, genetic confounding fully accounted for the screen time-internalizing problems relationship (Figure 3B, right panel; Table S5).

In sensitivity analyses using models with only outcome genetic information suggested larger estimates of genetic confounding (Figure S5). In another sensitivity analyses adjusting for both genetic and socioeconomic factors (SES; family income and highest parental education), we found similar percentages of genetic confounding effects, though the absolute effect magnitudes were reduced (Figure S6). In sensitivity analyses using parent-report screen time, which may be impacted by shared reporter bias, we found less obvious genetic confounding effect in the relation between screen time and attention problems (45.8%) or internalizing problems (19.5%) in the model using PRS and hSNP (Figure S7).

Finally, in sensitivity analyses testing alternative plausible values of hSNP for screen time and psychiatric problems, the genetic confounding effect completely explained the association between screen time and attention problems when the attention problem hSNP was set to 0.24 (assuming the screen time hSNP was 0.077; Figure S8A). Similarly, the residual association between screen time and internalizing problems disappeared when the hSNP of internalizing problems was set to 0.19 (Figure S8B).

## Discussion

This large population-based study of screen time and child psychiatric problems in preadolescents suggests that genetic confounding, if modelled with genetic information from PRSs and heritability may account for a substantial portion of the phenotypic association between screen time and child psychiatric problems. Our study entails several findings. First, increased child screen time was associated with more psychiatric problems, consistent with prior research in the ABCD study.^38^ This association was partially explained by sociodemographic factors and maternal psychopathology but largely remained after the adjustments. Second, we found specificity in associations between PRSs and their corresponding traits, e.g., the television time PRS was more strongly associated with child screen time than with other traits. However, we also found associations between PRSs and other traits, e.g., the television time PRS was associated with both attention and internalizing problems, suggesting horizontal pleiotropy of the genetic variants and thus possible genetic confounding, i.e., a genetic predisposition that could lead to more screen time and psychiatric problems.

This study aimed to quantify genetic confounding in the relationship between child screen time and psychiatric problems using a novel method, Gsens,^21^ that integrates information from both PRS and heritability estimates (either hSNP or htwin). The association between screen time and attention problems was strongly confounded by genetic factors. In contrast, the relationship between screen time and internalizing problems was moderately genetically confounded: some residual association remained when using hSNP, although none remained when using htwin. Importantly, this residual association encompasses both the direct effect of screen time on psychiatric problems and residual confounding by environmental factors (e.g., parenting practices that may independently impact both screen time and internalizing problems). Previous twin studies also similarly conclude that some lifestyle-behavior associations may be largely explained by genetic confounding.^39-41^

Our genetic confounding estimates highly depend on the magnitudes of heritability estimates. More importantly, hSNP and htwin estimates in our analytic sample were comparable with those of previous studies, despite differences in sample population, genetics quality control thresholds, and measurements (Table 2). With some fluctuations in the heritability estimates, the genetic confounding should be interpreted with caution. Although our study provides a range for potential genetic confounding based on hSNP and htwin, the true magnitude of genetic confounding may be less than the lower bound if hSNP was overestimated. Thus, the sensitivity analyses with a range of potential hSNP values further contextualize our findings.

Some socioeconomic status measures may partly index genetic factors. For example, adjusting for maternal psychopathology reduced the primary phenotypic associations possibly because mothers may transmit genetic variations related to both child screen time and psychiatric problems. Furthermore, the proportion of genetic confounding remained comparable even after additionally adjusting our models for SES. This suggests that adjusting for SES indeed captured some genetic confounding in the relationships between screen time and psychiatric problems, consistent with previous findings that multiple genes are related to both income and mental health.^42,43^

Our results highlight the importance of considering genetic factors in socio-behavioral research. Considering genetic influences helps us better understand complex causal relationships. Many policymakers and scientists view child screen time as an important modifiable risk factor. However, if genetic factors account for a large part of the observed relationship between screen time and mental health, interventions restricting child screen time could be less effective in preventing child attention and internalizing problems than expected,^44^ consistent with similar findings on genetic confounding between social media use on mental health.^45^ This does not suggest that parents should adopt a lenient attitude towards children using electronic devices excessively, as more screen time could be related with other risks, such as less physical or academic activities.

Our study has some limitations. First, the Gsens method assumes no gene-environment interactions. Second, we cannot rule out reverse causality even though we assessed the outcome at one-year follow-up. It is possible that children with a genetic vulnerability for ADHD are more likely to be exposed to more screen time, as a previous study discussed.^46^ However, if this were true, the direction of the association between screen time and attention problems might be reversed, but the conclusion regarding genetic confounding would not change. Third, we restricted the population to individuals of European ancestry, as most GWAS studies included European samples only. Furthermore, the PRSs in our analyses were derived from adult GWAS studies, which may make PRS-based genetic confounding estimates in children less precise.

In summary, this study suggests genetic confounding may account for much of the association between child screen time and attention problems, and part of the association between screen time and internalizing problems. These results demonstrate the importance of considering genetic confounding utilizing both PRSs and heritability information in socio-behavioral studies of modifiable factors for youth mental health.

## Data Availability

All data produced in the present study are available upon reasonable request to the authors

## Data sharing statement

Data for the ABCD Study are already open and available in the NIMH Data Archive (NDA) (nda.nih.gov) to eligible researchers within NIH-verified institutions. Data can be accessed following a data request to the NIH data access committee (https://nda.nih.gov/), which should include information on the planned topic of study. Data use should be in line with the NDA Data Use Certification.

