## Supplemental file for "The associations of child screen time with psychiatric problems: the role of genetic confounding"

### Supplementary Method

#### Genotyping

We restricted the genotyping to autosomes and removed duplicated SNPs and monomorphic SNPs. To identify European ancestry population, we found common SNPs between study sample and 1kg sample, with minor allele frequency (MAF) >0.05 and call rate >0.98 in both samples, then merged study sample with 1kg sample on the common high-quality SNPs extracted above, and removed strand ambiguous SNPs, indels and long-range LD regions. After LD pruning, an initial PCA on the merged sample were conducted. Compared with extracted self-reported ancestry, we identify 503 study samples who are self-reported White/Caucasian and have the shortest Euclidean distance to the center of 1kg EUR samples in the PC1-PC2 space. Then, we calculated MAF in the 503 EUR study samples and 503 1kg EUR samples and removed SNPs that have MAF difference >0.05 from the pruned set of SNPs (in order to remove SNPs that have potential genotyping errors). After the re-calculation of PCs using the updated set of pruned SNPs in the merged sample, population were assigned using random forest to extract EUR samples with predicted probability > 0.8. We kept common SNPs between study sample and 1kg sample and removed SNPs that have MAF difference > 0.05 between EUR study sample and 1kg EUR sample. We also removed samples that failed sex and heterozygosity check. After imputation from Michigan Imputation Server (reference panel HRC), we extracted a list of SNPs with MAF > 0.01, imputation Rsq > 0.8, SNP-level call rate > 0.98, and Hardy–Weinberg equilibrium p > 1e-10.

#### Child screen time

Child screen time information was collected at baseline with a 14-question Screen Time Questionnaire (STQ) completed by the children, providing self-report measures of screen time use, divided by weekdays and weekends. The questionnaire asks how many hours per weekday/weekend day the child uses different types of screen-based media, with responses ranging from “0 h” (0) to “4 + h” (4). The STQ assesses screen time use for six different forms of recreational media use: television shows and movies, videos, video games, texting, social media, and video chat. The total amount of time spent on screens on an individual weekday or weekend day is a composite across all six forms of media types. In sensitivity analysis and for comparison, we also used a shorter parent-report STQ. This shorter version assesses only the child’s total screen time on weekdays and weekend days in hours without specified subtypes of screen time. Daily screen time averaged across both weekdays and weekends was used in our analysis. In our analysis, we standardized both child-rated and parent-rated screen time to mean 0 and standard deviation (SD) 1 for better comparisons.

**Covariates**

Family income and parental education were considered as additional potential confounders in the associations between screen time and child psychiatric problems. Family annual income was categorized into 10 levels, from less than $5000 to more than $200,000. Parental education was evaluated as a categorical variable ranging from 0 representing ‘never attended school’ to 21 representing a doctoral degree (the highest attainment). We also additionally included maternal psychopathology as potential confounder to examine potential bias from using the parental informants for exposure and outcome, i.e., parents, on both child screen time and psychiatric problems. Meanwhile, Maternal psychopathology was included as a covariate to reduce the bias from using a parental informant for exposure and outcome. Maternal psychopathology was evaluated with continuous sum scores from the Adult Self Report questionnaire.[1]

**Heritability**

The heritability estimates of child-reported (or parent-reported) child screen time, attention problem raw scores, and internalizing problem raw scores were estimated using the largest available sample of unrelated individuals from ABCD study with European ancestry. Screen time and psychiatric domain scores were normalized prior to deriving the heritability estimates to fulfill the normal distribution assumption using GREML.

To conduct the identiy by descent (IBD) segments, we used the cut-off proportion of loci with 0 allele shared by descent (Z0) less than 0.1 and proportion of loci with 1 allele shared by descent (Z1) less than 0.1 to identify monozygotic twins and the cut-off Z0 larger than 0.125 and Z0 less than 0.375 to identify dizygotic twins.

Supplementary Figure 1 Genetic confounding sensitivity (Gsens) analysis framework.

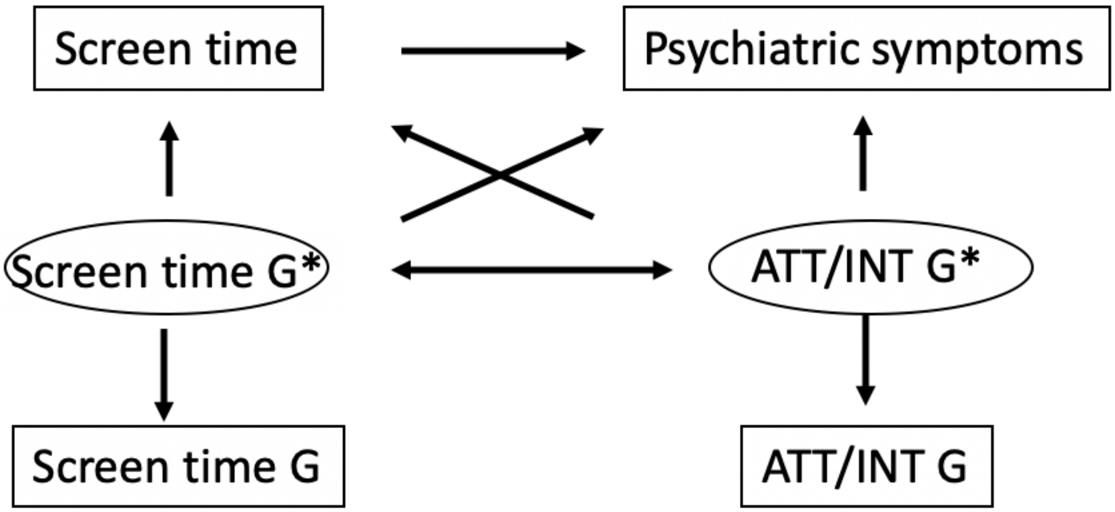

Figure S2: Comparison of associations between polygenic scores (PRS) and child-rated screen time versus child-rated television time only (N=4262).

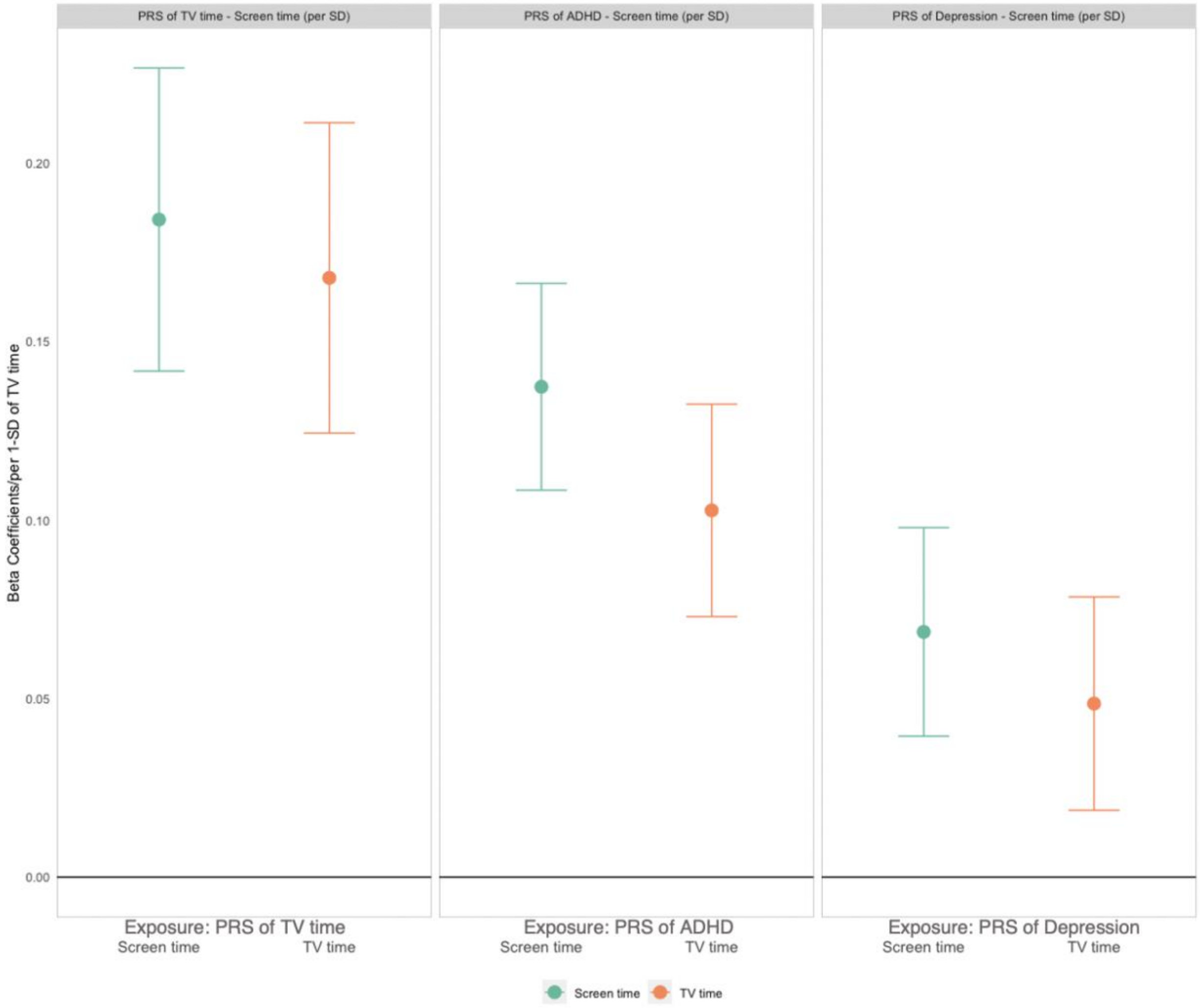

Associations adjusted for age, sex, site, and top 10 PCs.

Noted: Polygenic risk scores, child-rated screen time and TV time were standardized to mean 0 and standardized deviation 1.

Table S1: Demographic distribution of genetically identified European unrelated individuals compared with self-identified European children.

| | Genetically identified European (included) Mean ± SD / N (%) | Self-identified European (excluded) Mean ± SD / N (%) | p-value |
|---|---|---|---|
| N | 4262 | 4090 | |
| Age, year | 9.93 ± 0.62 | 9.90 ± 0.64 | 0.02 |
| Sex | | | |
| Female, % | 1993 (46.8) | 1949 (47.7) | 0.43 |
| Male, % | 2269 (53.2) | 2141 (52.3) | |
| Family annual income | | | |
| Less than $50,000, % | 510 (12.0) | 929 (22.7) | <0.001 |
| $50,000 - $100,000, % | 1326 (31.1) | 559 (13.7) | <0.001 |
| More than $100,000, % | 2426 (56.9) | 2602 (63.6) | <0.001 |
| Parent education level (degree) | | | |
| High School graduation, % | 939 (22.0) | 1553 (38.0) | <0.001 |
| Bachelor’s Degree, % | 1323 (31.0) | 1113 (27.2) | <0.001 |
| Graduate Degree or above, % | 2000 (46.9) | 1419 (24.7) | <0.001 |
| Maternal psychopathology, score | 20.81 ± 16.54 | 21.28 ± 17.64 | 0.21 |
| Screen time (child-rated) at baseline, hour | 3.18 ± 2.55 | 3.58 ± 2.78 | <0.001 |
| Screen time (parent-rated) at baseline*, hour | 1.16 ± 0.60 | 1.25 ± 0.71 | <0.001 |
| Attention problems at year 1 follow-up, score | 2.88 ± 3.42 | 3.43 ± 2.84 | < 0.001 |
| Internalizing problems at year 1 follow-up, score | 5.40 ± 5.60 | 5.28 ± 5.63 | < 0.001 |

Note: parent reported screen time questionnaire was less detailed than the child reported one, only asking for overall screen time in average.

P-values were calculated with t-test for continuous variables and z-test for categorical variables.

Figure S3: Comparison of associations between child-rated versus parent-rated reported screen time and psychiatric symptoms (N=4262).

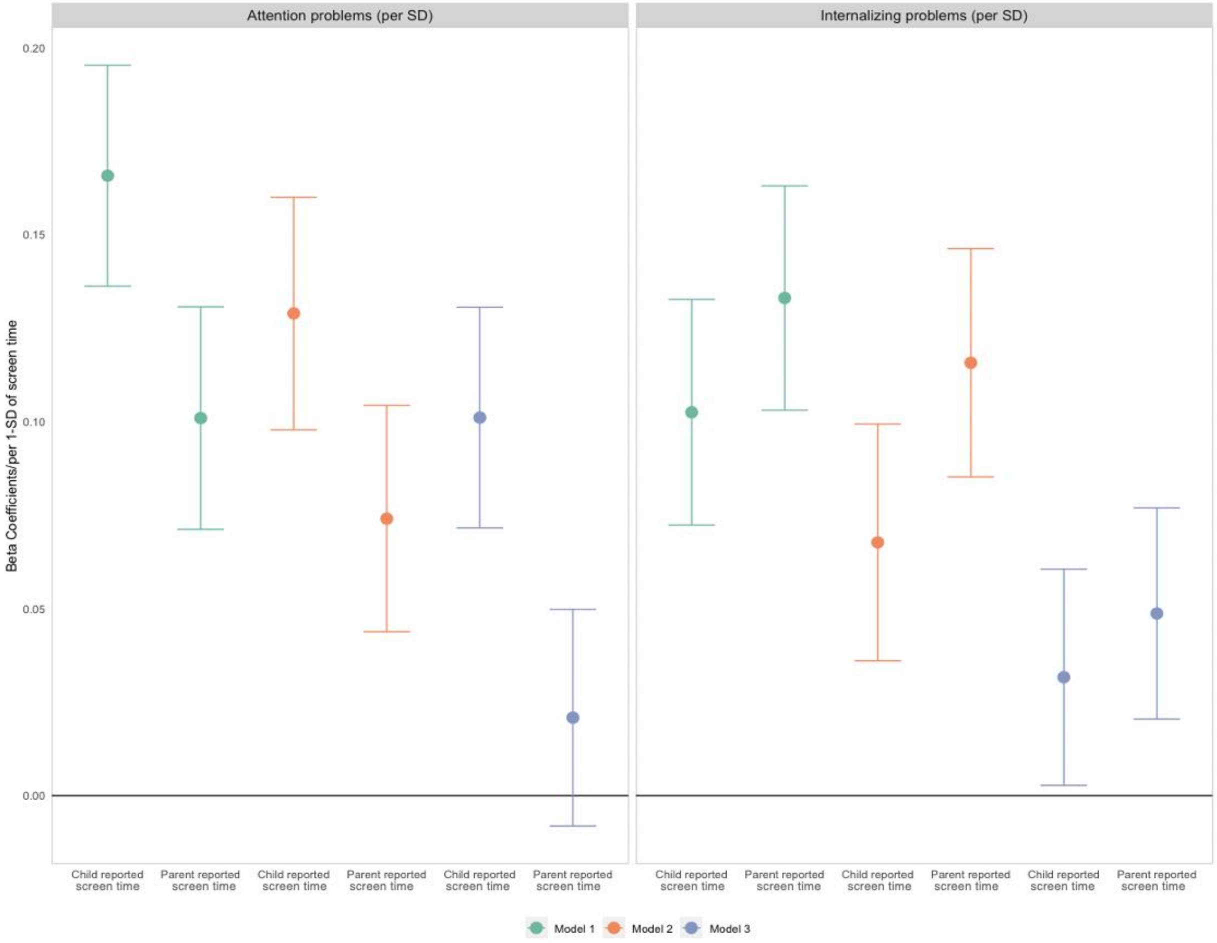

Model 1 adjusted for sex and age.

Model 2 adjusted for sex, age, family income, parental education.

Model 3 additionally adjusted for maternal psychopathology.

Note: Both child-reported and parent-reported screen time were standardized to mean 0 and standard deviation 1. Psychiatric symptoms include parent-reported attention and internalizing problems, standardized to mean 0 and standard deviation 1.

Table S2: Associations of child screen time with attention and internalizing problems.

(supplemented with Figure S2)

| Beta | SE | P value | 95% CI | Model |
|---|---|---|---|---|
| Child-rated screen time and Attention problems, 1-SD score/1-SD of screen time | | | | |
| 0.17 | 0.015 | 0.000 | 0.14, 0.20 | Model 1 |
| 0.13 | 0.016 | 0.000 | 0.10, 0.16 | Model 2 |
| 0.10 | 0.015 | 0.000 | 0.07, 0.13 | Model 3 |
| Child-rated screen time and Internalizing problems, 1-SD score/1-SD of screen time | | | | |
| 0.10 | 0.015 | 0.000 | 0.07, 0.13 | Model 1 |
| 0.07 | 0.016 | 0.000 | 0.04, 0.10 | Model 2 |
| 0.03 | 0.015 | 0.032 | 0.003, 0.06 | Model 3 |
| Parent-rated screen time and Attention problems, 1-SD score/1-SD of screen time | | | | |
| 0.10 | 0.015 | 0.000 | 0.07, 0.13 | Model 1 |
| 0.07 | 0.015 | 0.000 | 0.04, 0.10 | Model 2 |
| 0.02 | 0.015 | 0.159 | -0.008, 0.05 | Model 3 |
| Parent-rated screen time and Internalizing problems, 1-SD score/1-SD of screen time | | | | |
| 0.13 | 0.015 | 0.000 | 0.10, 0.16 | Model 1 |
| 0.12 | 0.016 | 0.000 | 0.09, 0.15 | Model 2 |
| 0.05 | 0.014 | 0.000 | 0.02, 0.08 | Model 3 |

Model 1 adjusted for sex and age.

Model 2 adjusted for sex, age, family income, parental education.

Model 3 additionally adjusted for maternal psychopathology.

Figure S4: Comparison of associations between polygenic scores (PRS) and child-rated versus parent-rated screen time (N=4262).

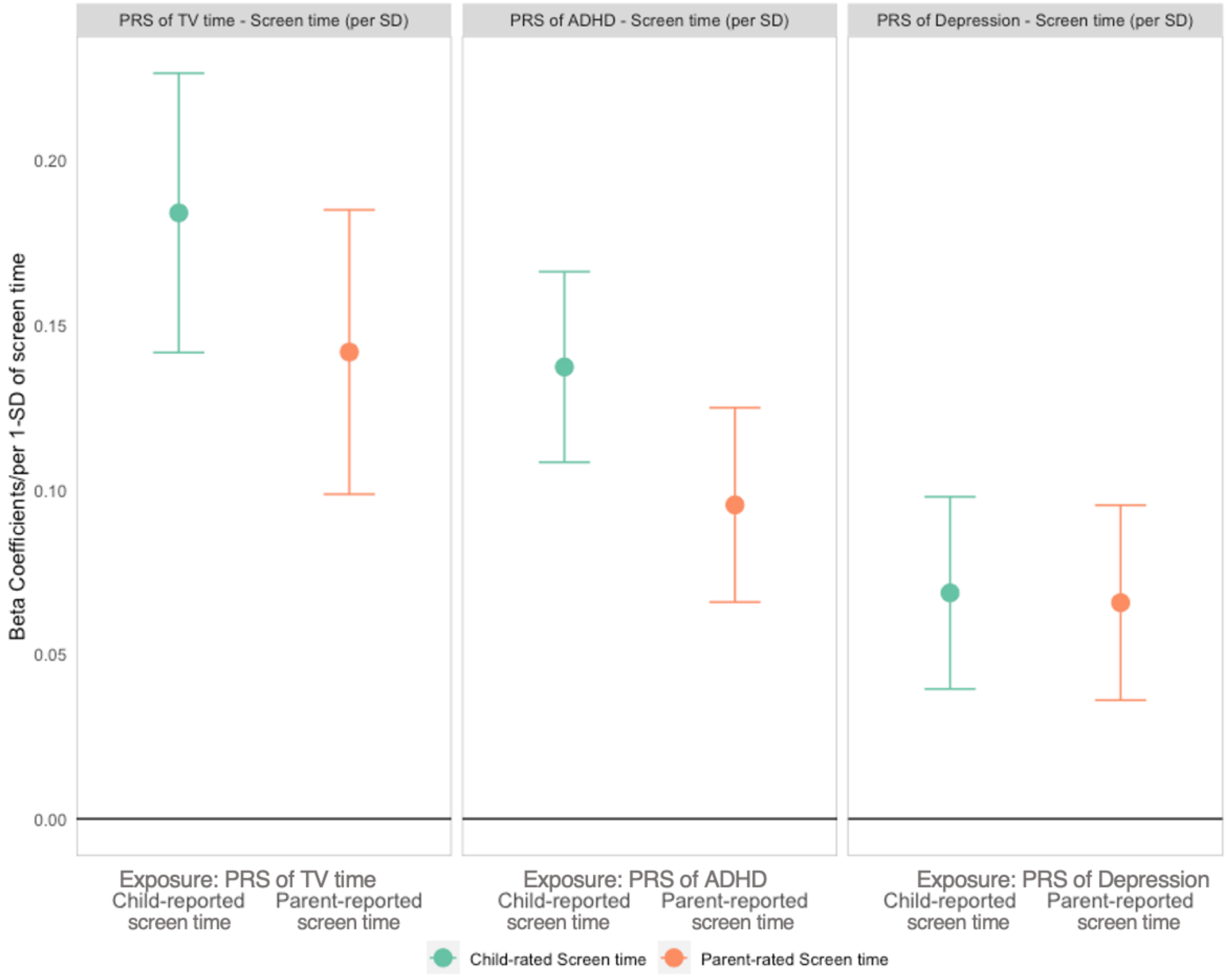

Associations adjusted for age, sex, site, and top 10 PCs.

Noted: Polygenic risk scores, child-rated screen time and parent-rated screen time were standardized to mean 0 and standardized deviation 1.

Table S3: Associations of polygenic scores with child-reported screen time, attention and internalizing problems (supplemented with Figure 2).

| Exposure | Outcome | Beta estimates (95% CI) | P value of Beta estimates | Explained variance |
|---|---|---|---|---|
| PRS of TV time | Screen time, per SD | 0.18 (0.14, 0.23) | 2.51e-17 | 1.55% |
| PRS of ADHD | | 0.14 (0.11, 0.17) | 2.33E-20 | 0.88% |
| PRS of depression | | 0.07 (0.04, 0.10) | 4.18E-06 | 0.40% |
| PRS of TV time | Attention problems, per SD | 0.09 (0.05, 0.13) | 4.62E-05 | 0.36% |
| PRS of ADHD | | 0.13 (0.10, 0.16) | 7.57E-17 | 1.56% |
| PRS of depression | | 0.07 (0.04, 0.10) | 1.21E-05 | 0.42% |
| PRS of TV time | Internalizing problems, per SD | 0.08 (0.04, 0.13) | 2.32E-04 | 0.29% |
| PRS of ADHD | | 0.05 (0.02, 0.08) | 1.23E-03 | 0.22% |
| PRS of depression | | 0.10 (0.07, 0.13) | 1.82E-10 | 0.92% |

Associations adjusted for age, sex, site, and top 10 PCs.

PRSs were all standardized to mean 0 and standard deviation 1

Table S4: Correlation matrix among child reported screen time, child psychiatric problems, and polygenic risk scores (N=4262).

| N=4262 | Child-rated Screen time | CBCL Attention problem score | CBCL Internalizing problem score | ADHD PRS | Depression PRS | TV time PRS |
|---|---|---|---|---|---|---|
| Parent-rated Screen time | 0.29 (0.25) | 0.10 (0.07) | 0.13 (0.11) | 0.10 (0.07) | 0.07 (0.05) | 0.10 (0.06) |
| Child-rated Screen time | 1 | 0.16 (0.13) | 0.10 (0.07) | 0.14 (0.10) | 0.07 (0.05) | 0.13 (0.08) |
| CBCL Attention problem score | | 1 | 0.47 | 0.13 | 0.07 | 0.06 |
| CBCL Internalizing problem score | | | 1 | 0.05 | 0.10 | 0.06 |
| ADHD PRS | | | | 1 | 0.21 | 0.19 |
| Depression PRS | | | | | 1 | 0.12 |
| TV time PRS | | | | | | 1 |

Note: PRS were adjusted for sex and top 10 PC, CBCL scores were additionally adjusted for sex, age and site, and screen time was additionally adjusted for sex, age, site. The estimated correlation in the parentheses used screen time residualzed on sex, age, site, family income and parental education. All variables were standardized to mean 0 and standardized deviation 1.

Figure S5: Association of child screen time (self-reported) with attention problems and internalizing problems: with different adjustments for genetic confounding using genetic information on the outcome (GsensY) (N=4262).

A

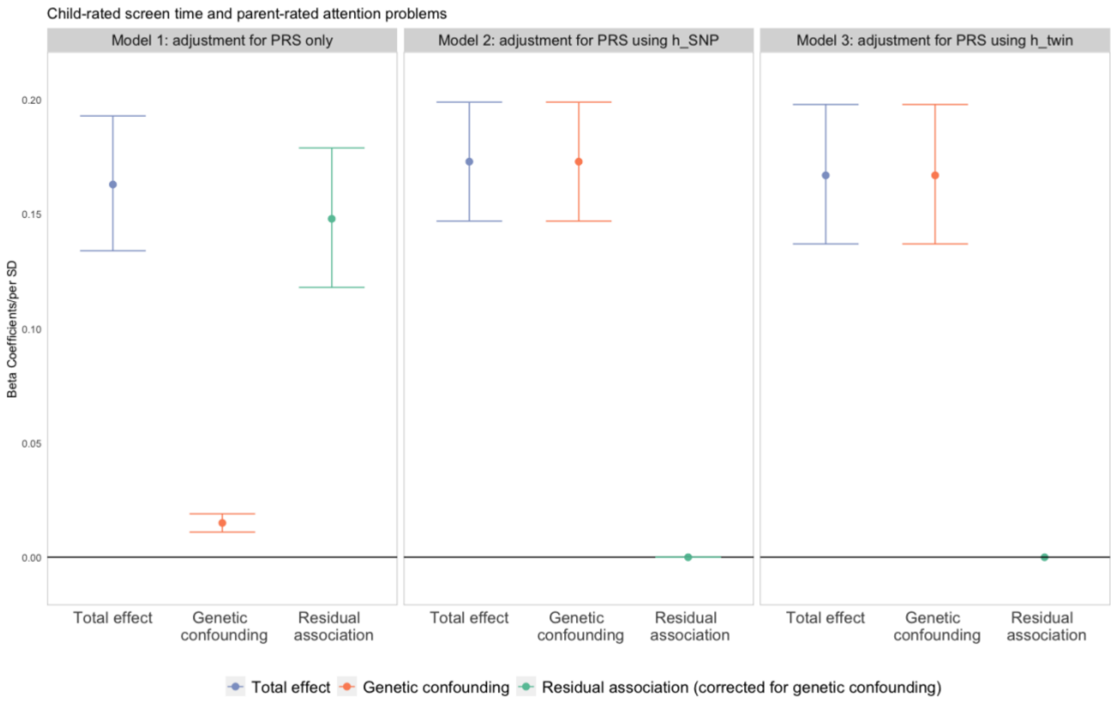

B

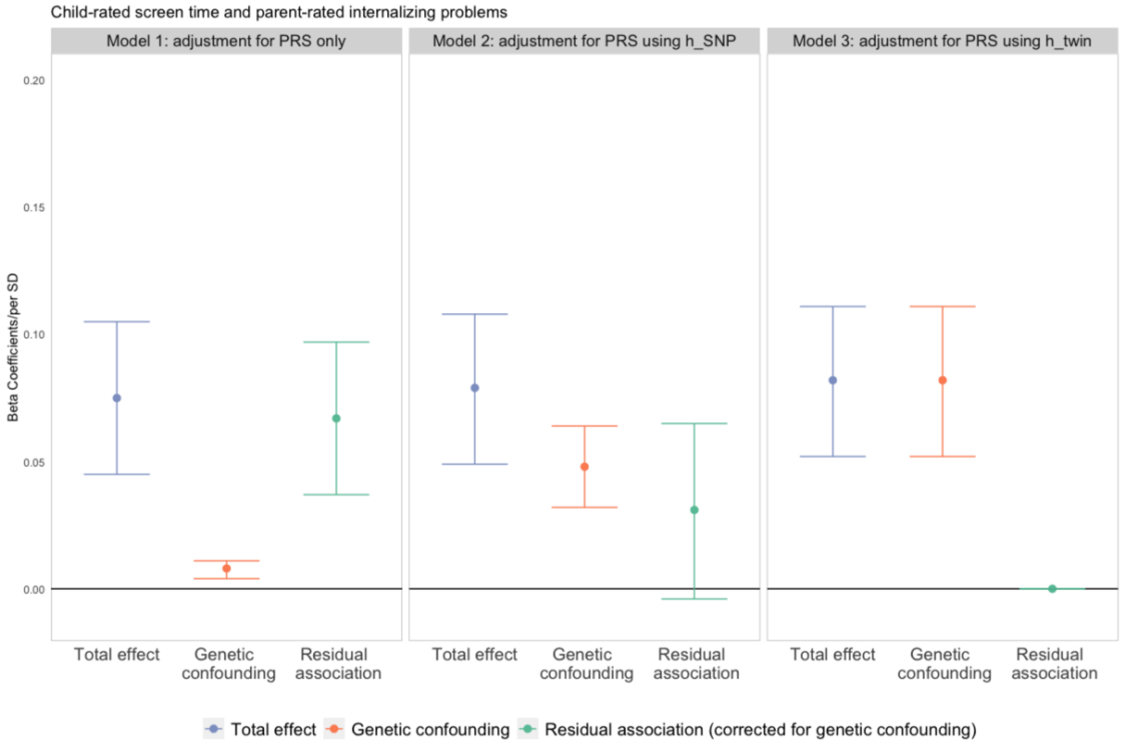

Model 1: adjusted for polygenic risk scores for outcomes only.
Model 2: adjusted for polygenic risk scores using SNP-based heritability ($h_{SNP}$) for outcomes.
Model 3: adjusted for polygenic risk scores using twin-based heritability ($h_{twin}$) for outcomes.

Figure S6: Association of child screen time (self-reported) with attention problems and internalizing problems: with different adjustments for genetic confounding and additionally residualizing on socioeconomic status (SES) (N=4262).

A

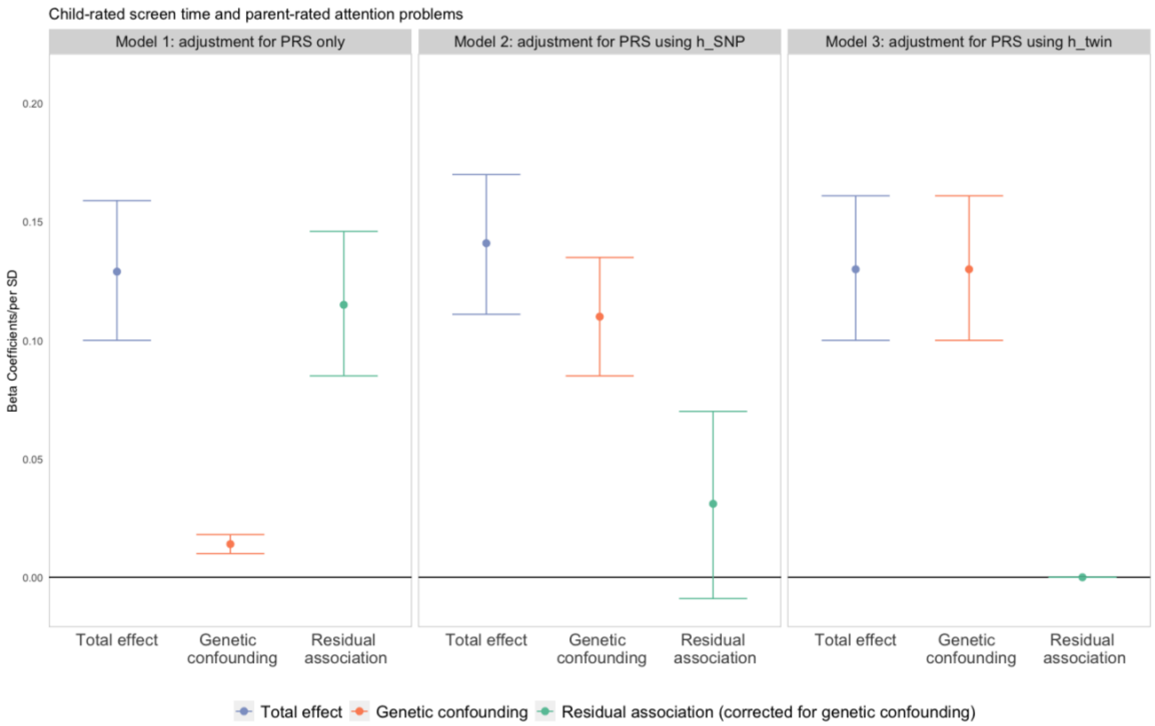

B

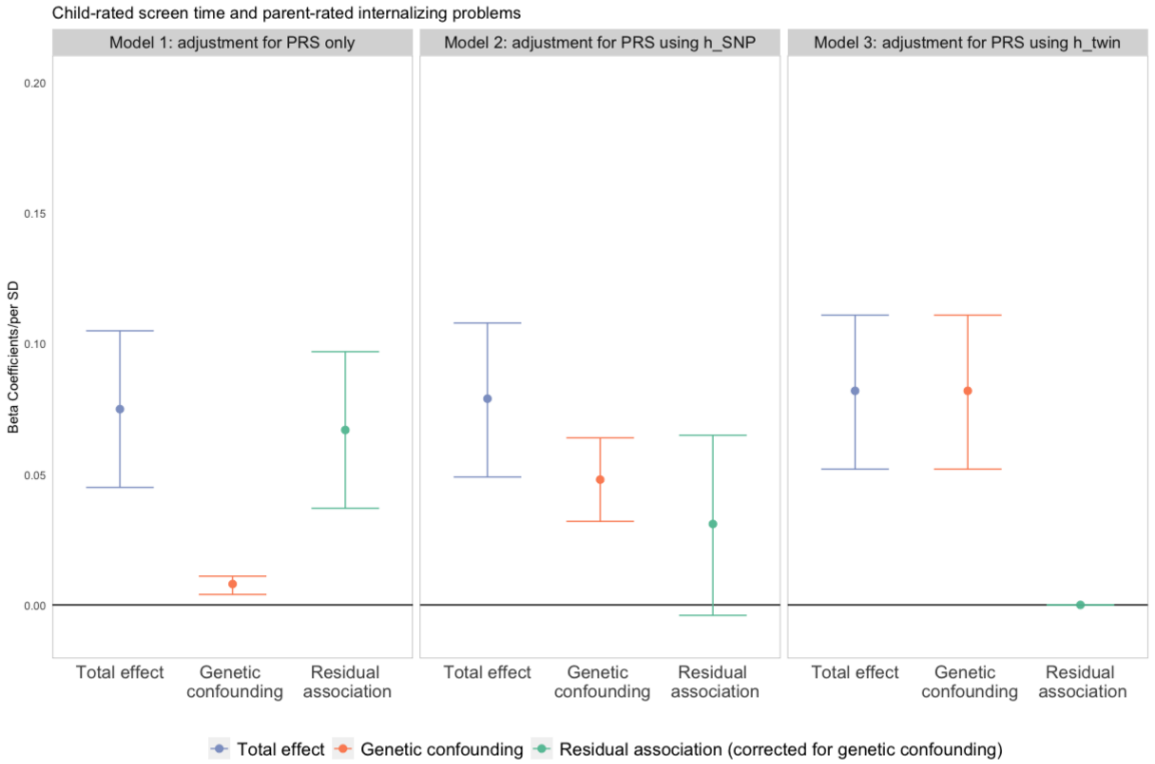

Model 1: adjusted for polygenic risk scores for both exposure and outcomes only.
Model 2: adjusted for polygenic risk scores using SNP-based heritability ($h_{SNP}$) for both exposure and outcomes.
Model 3: adjusted for polygenic risk scores using twin-based heritability ($h_{twin}$) for both exposure and outcomes.

Figure S7: Association of parent-rated child screen time with attention problems and internalizing problems: with different adjustments for genetic confounding (N=4262).

A

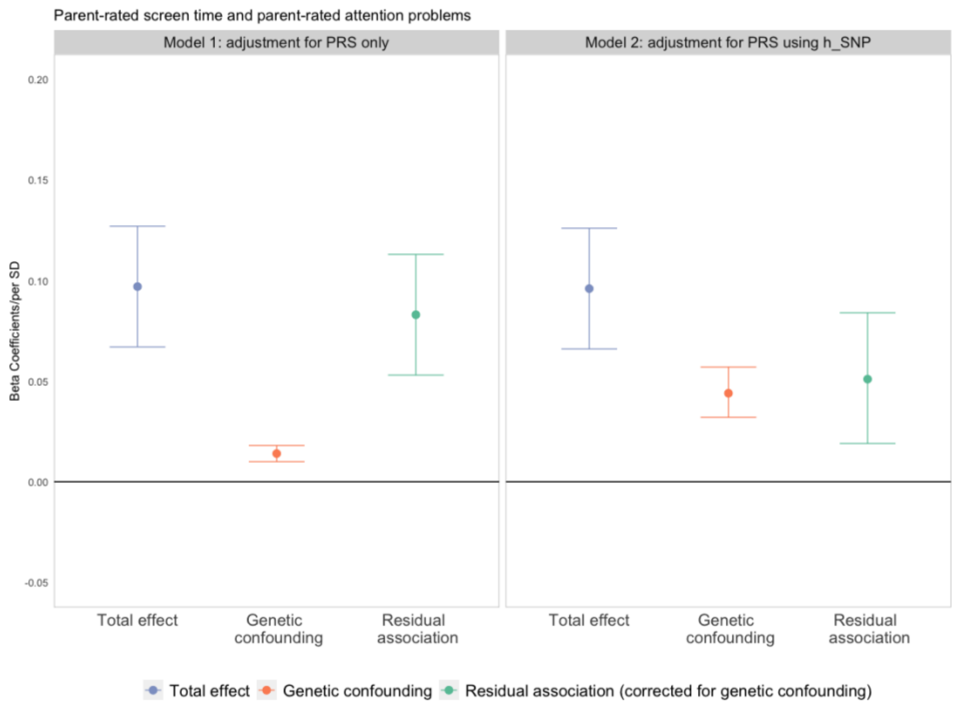

B

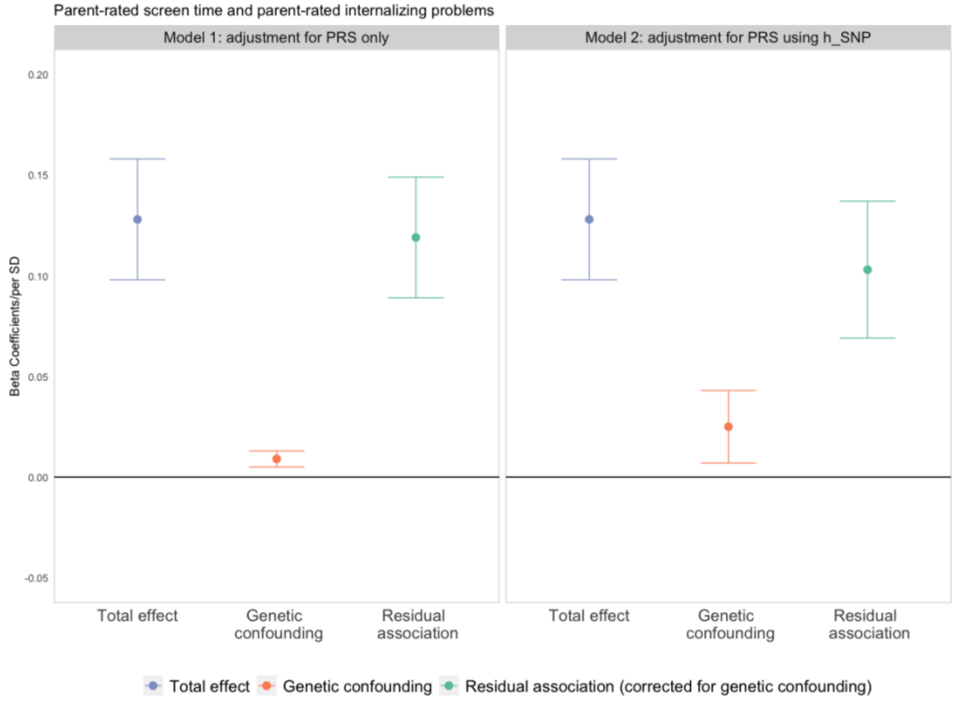

Model 1: adjusted for polygenic risk scores for both exposure and outcomes only.
Model 2: adjusted for polygenic risk scores using SNP-based heritability ($h_{SNP}$) for both exposure and outcomes.
Note: Because we could not obtain a reliable twin-based heritability estimate for parent-rated screen time, model 3 is not included in our analysis here.

Figure S8 Sensitivity analyses of genetic confounding using different SNP-based heritability simulated values of ADHD in two scenarios of screen time heritabilities.

A. Attention problems

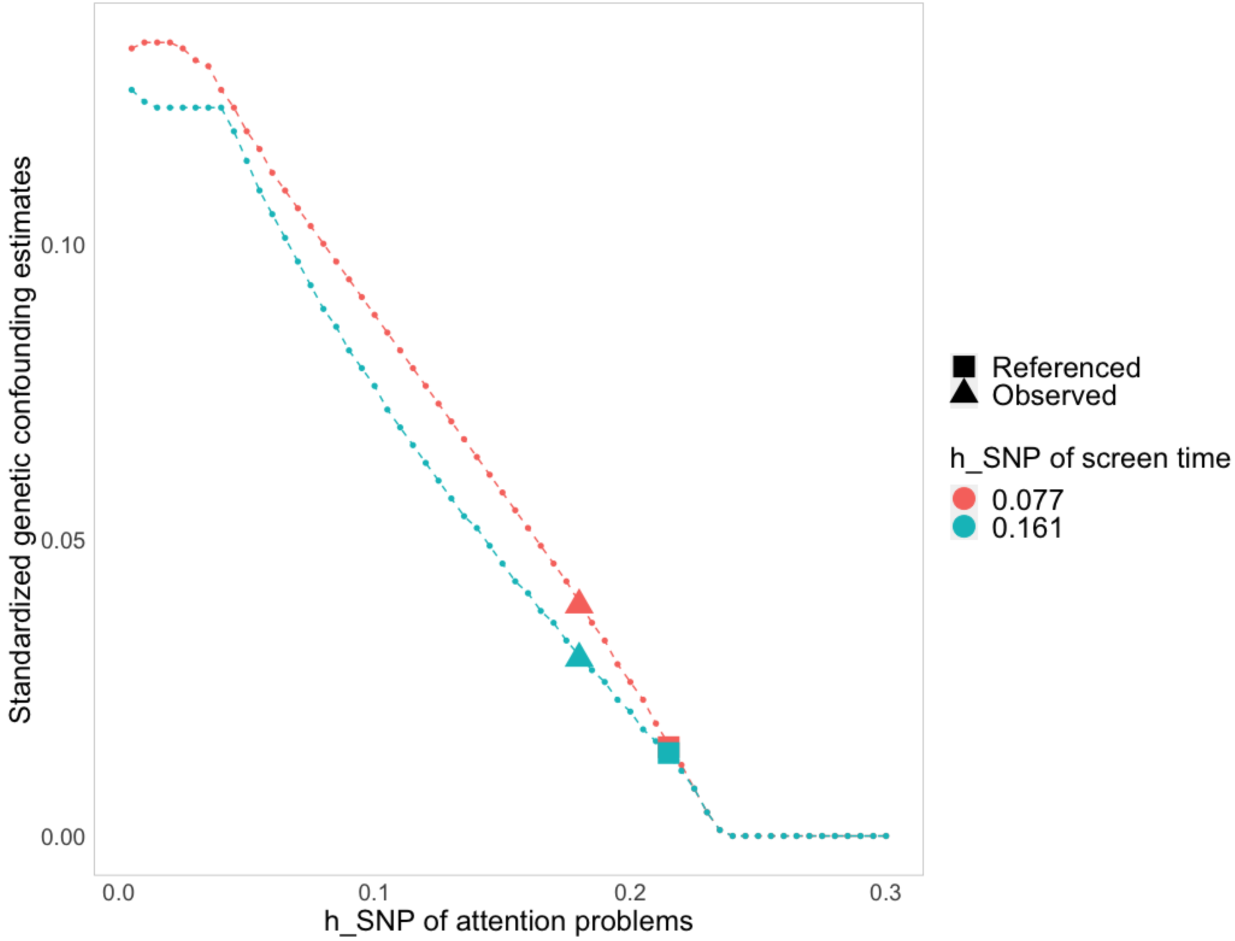

Note: Television time in adults was used as the phenotype in the reference of screen time heritability estimates (h2=0.16, green line). Clinically diagnosed ADHD cases and controls were used in the reference of attention heritability (showed as squares, h2=0.216). The triangle showed the observed h2 of attention problems (0.15) in our study sample and the circles showed the simulated possible h2 estimates of attention problems.

B. Internalizing problems

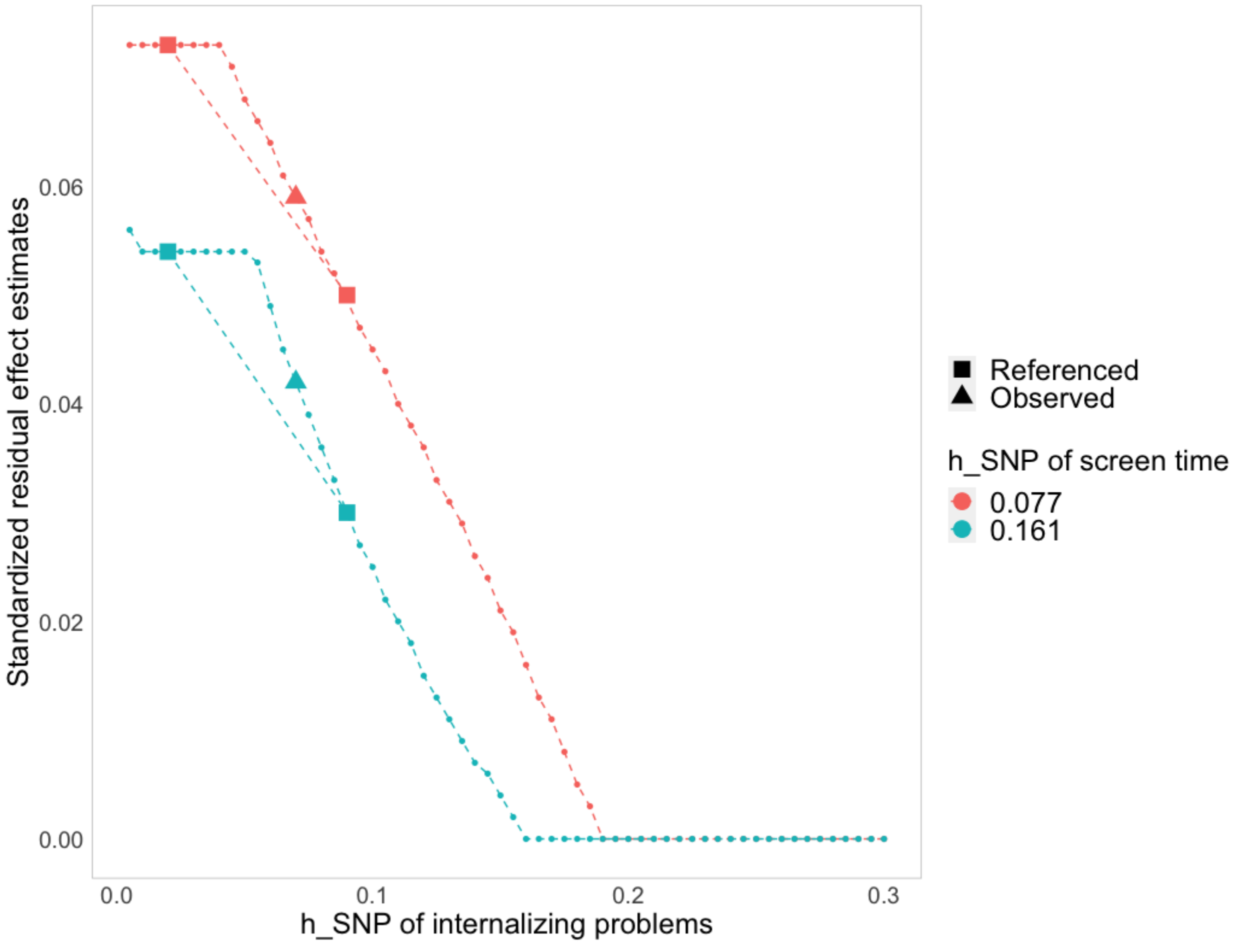

The Moods and Feelings Questionnaire (MFQ) reported by parents at 12-year-old was used as the phenotype in the reference (showed as squares, h2=0.017). The triangle showed the observed h2 of internalizing problems (0.036) in our study sample and the circles showed the simulated possible h2 estimates of internalizing problems.

Table S5: Association of screen time with attention and internalizing problems: with different adjustments for genetic confounding.

| | | Beta Estimates | 95% CI |
|---|---|---|---|
| Child-rated screen time - attention problems | | | |
| PRS only | Residual effect | 0.146 | 0.115, 0.176 |
| | Genetic confounding | 0.017 | 0.013, 0.022 |
| PRS using h2 | Residual effect | 0.038 | -0.009, 0.085 |
| | Genetic confounding | 0.139 | 0.104, 0.174 |
| PRS using H2 | Residual effect | 0 | NA |
| | Genetic confounding | 0.167 | 0.136, 0.197 |
| Child-rated screen time - internalizing problems | | | |
| PRS only | Residual effect | 0.090 | 0.060, 0.121 |
| | Genetic confounding | 0.010 | 0.006, 0.015 |
| PRS using h2 | Residual effect | 0.059 | 0.023, 0.095 |
| | Genetic confounding | 0.044 | 0.024, 0.063 |
| PRS using H2 | Residual effect | 0 | NA |
| | Genetic confounding | 0.111 | 0.082, 0.141 |
| Child-rated screen time - attention problems adjusting for SES | | | |
| PRS only | Residual effect | 0.115 | 0.085, 0.146 |
| | Genetic confounding | 0.014 | 0.010, 0.018 |
| PRS using h2 | Residual effect | 0.031 | -0.009, 0.070 |
| | Genetic confounding | 0.110 | 0.085, 0.135 |
| PRS using H2 | Residual effect | 0 | NA |
| | Genetic confounding | 0.130 | 0.100, 0.161 |
| Child-rated screen time - internalizing problems adjusting for SES | | | |
| PRS only | Residual effect | 0.067 | 0.037, 0.097 |
| | Genetic confounding | 0.008 | 0.004, 0.011 |
| PRS using h2 | Residual effect | 0.031 | -0.004, 0.065 |
| | Genetic confounding | 0.048 | 0.032, 0.064 |
| PRS using H2 | Residual effect | 0 | NA |
| | Genetic confounding | 0.082 | 0.052, 0.111 |

SES: socioeconomic status.
